# Adherence Monitoring Methods to Measure Virological Failure in People Living with HIV on Long-Term Antiretroviral Therapy in Uganda

**DOI:** 10.1101/2022.05.11.22274977

**Authors:** Stephen Okoboi, Joseph Musaazi, Rachel King, Sheri A. Lippman, Andrew Kambugu, Andrew Mujugira, Jonathan Izudi, Rosalind Parkes-Ratanshi, Agnes N. Kiragga, Barbara Castelnuovo

## Abstract

Appointment keeping and self-report within 7-day or and 30-days recall periods are non-objective measures of antiretroviral treatment (ART) adherence. We assessed incidence of virological failure (VF), predictive performance and associations of these adherence measures with VF among adults on long-term ART. Data for persons initiated on ART between April 2004 and April 2005, enrolled in a long-term ART cohort at 10-years on ART (baseline) and followed until December 2021 was analyzed. VF was defined as two consecutives viral loads ≥1000 copies/ml at least within 3-months after enhanced adherence counselling. We estimated VF incidence using Kaplan-Meier and Cox-proportional hazards regression for associations between each adherence measure (analyzed as time-dependent annual values) and VF. The predictive performance of appointment keeping and self-reporting for identifying VF was assessed using receiver operating characteristic curves and reported as area under the curve (AUC). We included 900 of 1,000 participants without VF at baseline: median age was 47 years (Interquartile range: 41-51), 60% were women and 88% were virally suppressed. ART adherence was ≥95% for all three adherence measures. Twenty-one VF cases were observed with an incidence rate of 4.37 per 1000 person-years and incidence risk of 2.4% (95% CI: 1.6%-3.7%) over the 5-years of follow-up. Only 30-day self-report measure was associated with lower risk of VF, adjusted hazard ratio (aHR)=0.14, 95% CI:0.05–0.37). Baseline CD4 count ≥200cells/ml was associated with lower VF for all adherence measures. The 30-day self-report measure demonstrated the highest predictive performance for VF (AUC=0.751) compared to appointment keeping (AUC=0.674), and 7-day self-report (AUC=0.687). The incidence of virological failure in this study cohort was low. Whilst 30-day self-report was predictive, appointment keeping and 7-day self-reported adherence measures had low predictive performance in identifying VF. Viral load monitoring remains the gold standard for adherence monitoring and confirming HIV treatment response.

## Introduction

The WHO public health approach for scaling up access to anti-retroviral therapy (ART) expanded following the availability of highly active anti-retroviral medication through standardized regimes and decentralized care in Low and Middle Income Countries (LMIC) (1). In this approach, standardized simplified ART regimens and decentralized treatment delivery enabled large numbers of people with HIV (PHW) to be initiated and followed-up on treatment through public and private sector health facilities. The approach is centred on “four Ss”, an acronym for when to Start drug treatment, Substitute for toxicity, Switch after treatment failure, and Stop to enable lower-level healthcare workers to deliver appropriate care (1).

In 2020, an estimated 17 million people were on ART in sub-Saharan Africa (SSA) (2); in Uganda, 1,275,000 million persons were estimated to be on ART in 2019 (3). The 2018 and 2020 HIV treatment guidelines in Uganda recommend ART adherence monitoring using non-objective measures including pill counts, appointment keeping, visual analogue scales, and self-reported pill use, used either individually or in combination (4,5). The use of these adherence measures encourages ART adherence discussions with patients and providing information about the risk of virological failure or to support daily tablet-taking behavior in settings where viral load testing is limited (6–8). However, in 2016, the Uganda Ministry of Health (MoH) HIV treatment guidelines recommended annual plasma HIV viral load for people on ART to monitor treatment effectiveness and identify individuals with detectable viral load (9). Annual viral load monitoring is recommended due to scarcity of resources in LMIC (10,11).

Sustained optimal adherence to ART ensures virological suppression, reduction in HIV-related morbidity and mortality, and prevents onward transmission (6,12,13), popularized by the Joint United Nations Programme on HIV/AIDS (UNAIDS) as “*Undetectable equals Untransmittable (U=U)* (14,15). However, previous studies report discrepancies in ART adherence thresholds used. And adherence measured as a categorical or a continuous constructs from patients or clinic reports affecting association and predictive performance between ART adherence measures and virological failure among PWH on ART (6,7,16– 18). The performance of ART adherence measures in predicting virological failure among adult PWH on long-term ART (i.e., ≥10 consecutive years of ART use), including the predictors for virological failure are not well described across ART programs in LMIC. Despite self-reporting being routinely used as an adherence proxy in clinical care, few studies have evaluated the incidence of virological failure, predictive performance, and associations of appointment keeping, self-report within 7-day or and 30-days recall periods with VF among adults on long-term ART.

Thus, this study aimed to describe the incidence of virological failure, compare the predictive performance of three ART adherence measures (7-days and 30-days self-reported pill use, and appointment keeping) and assess factors associated with virological failure among PWH on long-term first-line ART.

## Methods

### Study design and setting

This study was conducted at the HIV Centre of Excellence at the Infectious Diseases Institute (IDI) located in the Mulago Teaching Hospital in Kampala, the capital city of Uganda. The IDI clinic is a large out-patient clinic that currently serves over 8,000 patients living with HIV in five municipalities in Kampala.

This was a secondary analysis of a longitudinal cohort data of patients enrolled in the Long-Term ART cohort. The ART Long-Term cohort is an observational cohort of 1,000 patients who had been on ART for at least 10 years and were enrolled between May 2014 and September 2015 to be followed up for an additional 10 years (19). Patients were eligible and enrolled in the cohort if they were ≥18-years, were willing to participate in the cohort visits and comply with the study procedures, and were in their 10th consecutive year of WHO standard ART at IDI regardless of the combination of drugs for first-line ART. Ten-year consecutive ART use was determined using data collected in the IDI electronic database, known as the Integrated Clinic Enterprise Application (ICEA). This is an in-house built system based on Microsoft.NET technologies (19). This interim analysis describes the first five years of follow-up.

### Data Collection

General medical history, physical examination, adherence to ART, and prescription of drugs were performed at enrolment and all study visits. Follow-up visits were scheduled once a year for 10-years. In addition to study visits, the participants attended the general clinic every 3-months to pick up their ART and concomitant medications. Antiretroviral drugs are prescribed according to the WHO guidelines; patients with two consecutive viral loads >1000 copies/ml after enhanced adherence counselling are considered for treatment switch. Enhanced adherence counselling is a targeted counselling offered to PWH on ART with non-suppressed viral load, done every month for at least 3-months before the next viral load test (5). At each study visit, real-time data entry into ICEA is performed by the respective providers (19). Laboratory results performed in the IDI Core Laboratory are automatically downloaded daily into the ICEA database. The questionnaires administered at each visit include basic demographic and epidemiological data, clinical history, adherence to ART, quality of life, and sexual behavior. Clinical data collected at each visit included vital signs and body weight, hematological and chemistry laboratory results, medications and ART regimen, and drug toxicities. All the data collected into ICEA are validated by a quality control and assurance officer who ensures that the data are complete and consistent.

### Adherence Measures

The primary outcome was virological failure defined as two consecutive plasma HIV RNA viral load measurements ≥1000 copies/ml at least within 3-months after receiving enhanced adherence counselling following the first viral load measurement. The exposure was ART adherence assessed using 3 different measures: self-reported pill use in the last 7 days, self-reported pill use in the last 30 days, and appointment keeping. The 30-day and 7-day self-report of pill use ART adherence measure was assessed on a scale of 1-100 by asking the patient to recall the numbers of missed doses in the last 30-days and 7-days and then calculating, what percentage of ART doses were taken. Good adherence was determined as having a score ≥95%. Appointment keeping was defined as returning for a scheduled cohort clinic visit appointment or within a 7-day window after a missed clinic visit. Questions were assessed throughout the entire 5-year follow-up period. Additional co-variates included age, sex, marital status, employment status, HIV disclosure status, household level of income, and body mass index.

We extracted cohort data from 10 to 15 years on ART follow-up (or enrollment in the cohort and five years of follow-up). When the required data were missing, patient charts were retrieved and reviewed to supplement the data in the databases. We extracted clinical data including ART start dates and regimens, socio-demographics at cohort enrolment, behavioral data, CD4 cell counts, plasma HIV viral load measurements for the follow-up period using or Roche COBAS® Ampli Prep. We also extracted data on deaths, transferred out, and lost to follow-up.

### Statistical analysis

Statistical analysis was performed using STATA 16.1 (StataCorp, College Station, Texas). We described cohort participants using frequencies and percentages for categorical variables and continuous variables using means and standard deviations and medians and interquartile ranges. Adherence measures were described using frequency and percentages across calendar year. Kaplan-Meier methods were used to estimate incidence risk and incidence rate of virological failure. Associations between virological failure and ART adherence was examined using Cox-proportional hazards regression (Cox-PH) models. ART adherence measures were entered into the model as time-dependent covariates measured at annual cohort visits. The Schoenfeld residuals test was used to assess for violation of the Cox –PH assumption. Three sensitivity analyses were performed, by refitting the model when: 1) missing values on covariates were imputed by multiple imputation using chained equations (MICE), 2) considering all censored patients i.e., deaths and losses to follow-up as virological failure (worst-case scenario), and 3) when considered as non-virological failure (best-case scenario). Performance of ART adherence measures - appointment keeping, 30-days and 7-days self-report of pill use for predicting virological failure was evaluated using receiver operating characteristic curve analysis. All hypothesis tests were performed as 2-tailed tests at a 5% significance level.

### Ethical approval

This study was approved by the Infectious Diseases Institute Research Ethics Committee (reference number; IDI REC-041/2021) and the Uganda National Council for Science and Technology (reference number; HS1896ES). The IDIREC committee granted a waiver of informed consent since secondary data were retrieved and analysed.

## Results

### Study Profile

We retrieved data for 1,000 PWH adults enrolled in the long-term ART cohort who started ART between April 2004 and April 2005 and were followed up until December 2021. Of the 1,000 participants, 100 (10%) had a viral load (VL) >1000 copies/ml documented at cohort enrolment and were therefore excluded from the study. Nine hundred participants were included in the analysis, of whom 10 had transferred to other health facilities, 45 were lost to follow-up and 41 had died before reaching 15 years on ART **(Fig 1)**.

**Fig 1:**
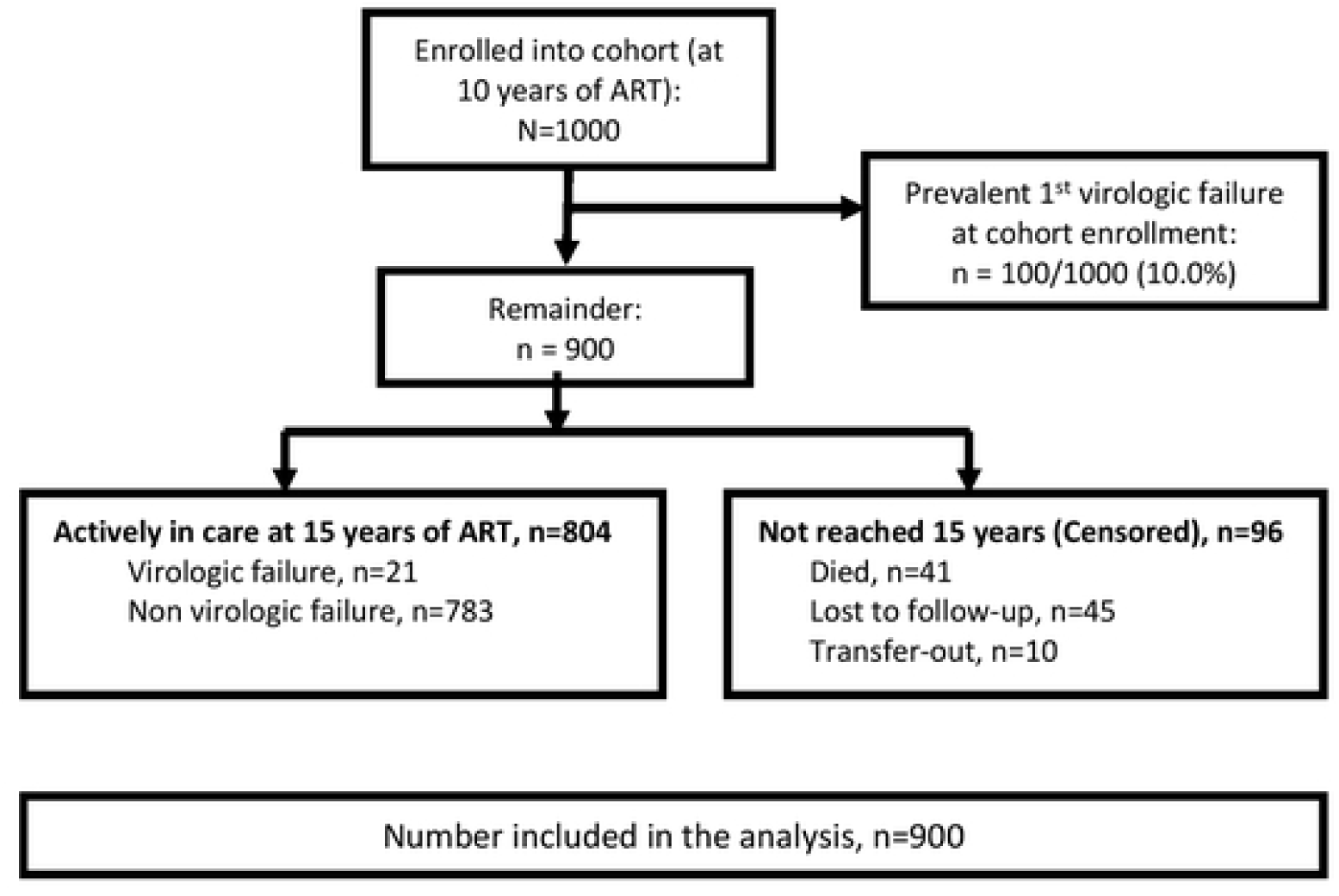
Cohort data flow diagram. Fig 1: Descript on of study participants analyzed

### Participant characteristics

#### Participants’ description

Of the 900 cohort participants analyzed, at cohort enrollment: the median age was 46 years (IQR 41-51); 59.8% were females, 82.1% were employed, 43.9% lived <1 US dollar per day, median body mass index (BMI) was 22.4 (IQR 19.8-25.4), 51.3% were married or cohabiting, and 88.4% had viral load <50 copies/ml (10 years on ART), (**Table 1)**.

**Table 1:** Baseline characteristics (at 10 years on ART)

footnote: SD denotes standard deviation, IQR interquartile range, BMI body mass 250 index. Missing values: BMI (n=42, 5%), Baseline VL (n=182, n=20%), CD4 count (n=81, 9%), 251 employed (n=5, 0.6%), household monthly income (n=49, 5%)
| **Variables** | **Statistics**<br>**N=900** |
| --- | --- |
| **Sex** |  |
| Male | 362 (40.2) |
| Female | 538 (59.8) |
| **Age (years) at cohort 2 registration** |  |
| Median (IQR) | 46.0 (41.0, 51.0) |
| **Age categories** |  |
| 28-44 | 388 (43.1%) |
| 45-54 | 368 (40.9%) |
| ≥55 | 144 (16.0%) |
| **BMI at baseline (cohort registration)** |  |
| Median (IQR) | 22.4 (19.8, 25.4) |
| **BMI <18kg/m<sup>2</sup>, N (%)** | 108 (12.6) |
| **VL at cohort 2 registration (c/ml)** |  |
| Median (IQR) | 20.0 (20.0, 20.0) |
| **Baseline VL categories (copies/ml), N (%)** |  |
| <50 | 635 (88.4) |
| ≥50 | 83 (11.6) |
| **CD4 at cohort 2 registration (cells/ml)** |  |
| Median (IQR) | 491.0 (347.0, 662.0) |
| **CD4 <200 cells/ml, N (%)** | 36 (4.4) |
| **Marital status, N (%)** |  |
| Single/Separated/Divorced/Widowed | 438 (48.7) |
| Married/Cohabiting | 462 (51.3) |
| **Employed, N (%)** |  |
| No | 160 (17.9) |
| Yes | 735 (82.1) |
| **Household monthly income (as <30\$ vs ≥30\$), N (%)** | |
| <30\$ per month | 374 (43.9) |
| ≥30\$ per month | 477 (56.1) |
| **Disclosure status, N (%)** |  |
| No | 830 (92.2) |
| Yes | 70 (7.8) |

The virological failure incidence rate was 4.37 (95% CI: 2.85 - 6.70) per 1000 person-years and the probability of virological failure was 2.4%, (95% CI: 1.6% - 3.7%) over 15 years, (**Fig 2)**. ART adherence was very high (≥95%) over the 7-calendar years studied on all adherence measures, except appointment keeping declined in 2019 and 2020 (42.0% and 72.7%, respectively) **(Table 2)**.

**Table 2:** ART Adherence measures (self-reported and appointment keeping during 2014 – 2020.

|  | 2014<br>(N = 137) | 2015<br>(N = 375) | 2016<br>(N = 968) | 2017<br>(N = 960) | 2018<br>(N = 940) | 2019<br>(N = 896) | 2020<br>(N = 343) |
| --- | --- | --- | --- | --- | --- | --- | --- |
| **7-day self-reported adherence** |  |  |  |  |  |  |  |
| No | 4 (2.9%) | 9 (2.4%) | 38 (3.9%) | 28 (2.9%) | 18 (1.9%) | 9 (1.0%) | 5 (1.5%) |
| Yes | 133 (97.1%) | 366 (97.6%) | 929 (96.1%) | 931 (97.1%) | 922 (98.1%) | 885 (99.0%) | 329 (98.5%) |
| **30-day self-reported adherence score $\geq 95\%$)** |  |  |  |  |  |  |  |
| No | 1 (0.7%) | 1 (0.3%) | 241 (26.1%) | 180 (18.8%) | 72 (7.7%) | 38 (4.3%) | 8 (2.5%) |
| Yes | 135 (99.3%) | 373 (99.7%) | 682 (73.9%) | 776 (81.2%) | 864 (92.3%) | 851 (95.7%) | 313 (97.5%) |
| **Appointment keeping adherence measure** |  |  |  |  |  |  |  |
| No | 5 (3.6%) | 23 (6.2%) | 65 (6.8%) | 46 (4.9%) | 64 (7.0%) | 403 (58.0%) | 83 (27.3%) |
| Yes | 132 (96.4%) | 346 (93.8%) | 891 (93.2%) | 896 (95.1%) | 849 (93.0%) | 292 (42.0%) | 221 (72.7%) |

**Missing data:** Self-reported (0,0,1,1,0,2,9 for 2014, 2015, 2016, 2017, 2018, 2019, 2020 respectively), Appointment keeping (0,6,12,18,27,201,39 for 2014, 2015, 2016, 2017, 2018, 2019, 2020 respectively), VAS (1,1,45,4,4,7,22 for 2014, 2015, 2016, 2017, 2018, 2019, 2020 respectively)

**Fig 2:**
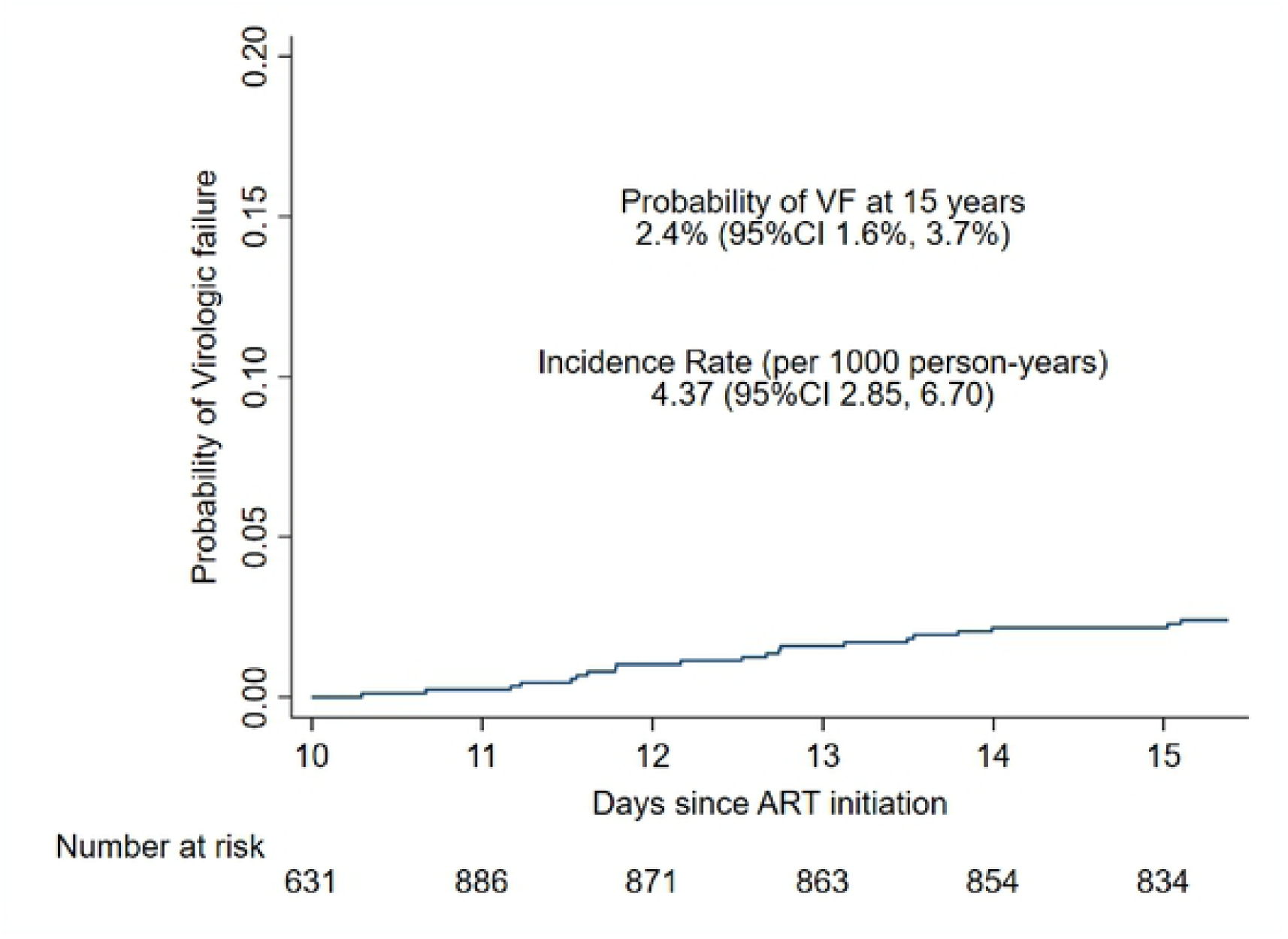
Probability and incidence rate of virological failure among persons on long-term. Kaplan-Meier indicates the probability of virologic failure and incidence rate at 15 years of follow-up among long-term persons on ART

### Associations between virologic failure and ART adherence measures

Table 3 shows that after adjusting for other patient factors, ART adherence assessed using 30-day self-report was associated with lower risk of virological failure (adjusted hazard ratio [AHR] 0.14; 95% CI: 0.05 - 1.76). However, the relationship was not significant when ART adherence was measured using 7-day self-report or appointment keeping (AHR 0.36; 95% CI: 0.05 – 2.75 and AHR 2.27; 95% CI: 0.27–18.83), respectively. Among other patient factors, only baseline CD4 ≥200 cells/ml was associated with lower risk of virological failure in models including all three adherence measures: 30-day self-report (AHR 0.24; 95% CI: 0.07–0.85), 7-day self-report (AHR 0.26; 95% CI: 0.08 – 0.91) and appointment keeping (AHR 0.22; 95% CI: 0.06 – 0.76). In sensitivity analyses, when imputing missing covariate data, the association between adherence and virological failure only remained for 30-day ART self-report (AHR 0.14; 95% CI: 0.05 – 0.35). There was no significant association with 7-day self-reported or appointment keeping measures **(Supplementary tables)**.

**Table 3:** Associations of ART adherence and virologic failure, adjusting for baseline and time-dependent patient characteristics.

| Factor | Model 1<br>30-day self-report ART<br>adherence measure<br>(as main exposure)<br>(n=800) |  | Model 2<br>7-day self-report ART<br>adherence measure<br>(as main exposure) (n=800) |  | Model 3<br>ART adherence measure<br>Appointment keeping (as main<br>exposure)<br>(n=795) |  |
| --- | --- | --- | --- | --- | --- | --- |
|  | aHR (95% CI) | P value | aHR (95% CI) | P value | aHR (95% CI) | P value |
| **ART adherence (time-updated)** |  |  |  |  |  |  |
| Non-adherent | 1 |  | 1 |  | 1 |  |
| Adherent | 0.14 (0.05 – 0.37) | <0.001 | 0.36 (0.05 – 2.75) | 0.325 | 2.27 (0.27 – 18.83) | 0.447 |
| **Other factors adjusted** |  |  |  |  |  |  |
| **Sex** |  |  |  |  |  |  |
| Male | 1 |  | 1 |  | 1 |  |
| Female | 2.29 (0.74 – 7.07) | 0.149 | 2.46 (0.81 – 7.50) | 0.133 | 2.84 (0.80 – 10.06) | 0.106 |
| **CD4 count at cohort registration (cells/mL)** |  |  |  |  |  |  |
| <200 | 1 |  | 1 |  | 1 |  |
| ≥200 | 0.24 (0.07 – 0.85) | 0.027 | 0.26 (0.08 – 0.91) | 0.035 | 0.22 (0.06 – 0.76) | 0.017 |
| **Employment status** |  |  |  |  |  |  |
| Unemployed | 1 |  | 1 |  | 1 |  |
| Employed | 0.57 (0.21 – 1.55) | 0.275 | 0.57 (0.22 – 1.53) | 0.267 | 0.60 (0.21 – 1.76) | 0.353 |

**Footnote:** ART adherence modelled as time-dependent covariates in all models. Analysis performed on complete cases for all covariates adjusted in the model: 11% (100/900) missing in model 1 and 2, whereas 12% (105/900) missing in model 3. **aHR** denotes adjusted hazard ratio from Cox proportional hazard regression models, **CI** denotes confidence interval. Apart from adherence (the main exposure), in the adjusted models we included only covariates with P value<0.2 at unadjusted model. Age groups, marital status and household monthly income had P values>0.2 in unadjusted Cox models, and thus were excluded from adjusted models.

### Adherence predictive performance of virologic failure

In the receiver operating characteristics curve (ROC) analysis for predictivity ability of adherence measures for virological failure, 30-day self-report best predicted virological failure (area under the curve [AUC] 0.751; 95% CI: 0.66 - 0.90) versus appointment keeping (AUC 0.674; 95% CI: 0.53 - 0. 81) and 7-day self-report (AUC 0.687; 95% CI: 0.51-0.82) (**Fig 3)**.

**Fig 3:**
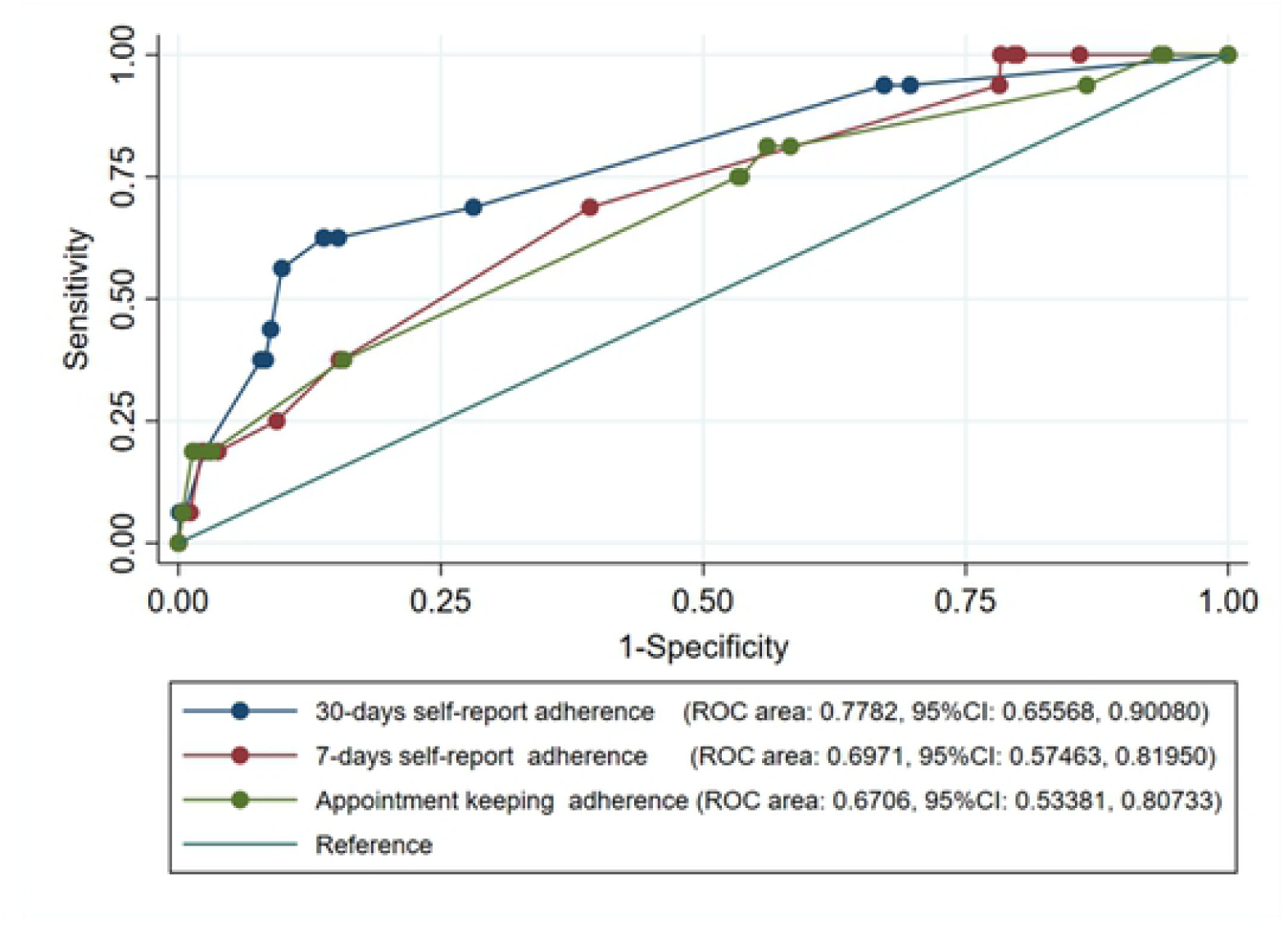
ROC curves for the three ART adherence measures in predicting virological failure. Fig 3: Predictivity ability of three adherence measures in predicating virological failure among long-term ART person

## Discussion

Overall, we found that unlike 30-day self-report, appointment keeping and 7-day self-reported adherence measures had low predictive performance in identifying virological failure. We also found a low incidence of virological failure among person living with HIV in this long-term ART study cohort. Our finding of low incidence of virological failure is consistent with an observational cohort analysis conducted in Uganda at The AIDS Support Organization among 3,340 persons who initiated ART from 2004 - 2009 and followed-up for a median of 5.7 years (IQR, 4.1 - 7.2 years) which found a low rate of virological failure among adult HIV patients on first-line antiretroviral therapy (20). Our study reports a low incidence of virological failure comparable to 7.4% reported among participants in the first Infectious Diseases Institute cohort followed up-to 10 years on ART (21). The lower incidence of virological failure observed in our study could be attributed to the longer duration on ART and the fixed dose once-daily therapy which was introduced around the time of cohort enrollment (19,22,23). Participants on long-term ART have received many ART adherence counselling sessions that should increase awareness about the importance of ART adherence and possibly drug side effects (21, 22). Furthermore, this is a survivor cohort of persons who have been on ART for at least ten years; there is evidence that shorter ART duration is conversely associated with increased risk of virological failure (23). This is due to the recent policy of universal test and treat, which increase the likelihood that patients with a new diagnosis of HIV tend to be unprepared to start ART as they have had limited psychosocial support due to rapid initiation (23,24). The issues of stigma, sero-status disclosure to people close to them and discrimination is another concern among people on ART as it increases non-adherence to treatment (23,24).

We found that baseline CD4 cells ≥200 cells/ml was associated with lower risk of developing virological failure in models using each of the three adherences measures. Our finding is similar to several studies both in developed and LMIC that have reported low baseline CD4 count have increased risk of developing virological failure (26). This finding is supported by other studies which recommended closer monitoring and ART adherence counselling for persons who commence ART with low CD4 count (26,27).

We also found that all the three adherence measures had low predictive performance in identifying virological failure. However, the 30-day self-report adherence measure was most able to predict virological failure. Our finding that 30-day self-report predicted virological failure is consistent with a study by Minyi et al., 2008 (28) who found that 1-month self-report ART adherence was more accurate in measuring ART adherence and predicating virological failure than 3-day or 7-day self-reported ART adherence (28–30). Other studies conducted in sub-Saharan Africa have found that self-report adherence measures have low predictive performance in detecting virological failure among participants on long and short-term ART (29). This could be because each of these adherence measures has inherent weakness such us their accuracy and precision due to recall and social desirability in different settings (25). Therefore, viral load monitoring as per WHO, remains the gold standard for identifying virological failure, monitoring ART adherence, and confirming treatment failure among people on ART even if its scale-up in resource-limited setting is hindered by financial and technical constraints (31).

To improve ART adherence, HIV care programs in LMIC should continue to educate people living with HIV on the importance of reporting accurate and consistent ART adherence, keeping dosing schedules, and explaining adverse effects. National ART guidelines should pay particular attention to monitoring virological failure and supporting ART adherence among persons who initiate ART with lower CD4 count. Monitoring viral load helps identify PWH on ART who have sustained long-term viral load suppression and is crucial in HIV prevention efforts given that national programs are promoting the UNAIDS slogan of *undetectable equals to untransmissible (U=U)*. As we disseminate and implement the UNAIDS policy, programs should integrate objective methods of measuring adherence like medication event monitoring systems, and biologic measures like point of care tenofovir testing that best predict virological failure.

The key strength of our study is the prospective data collection design among long-term ART persons, large sample size, long duration of follow-up, and objective ascertainment of virological failure. However, our study has limitations. These findings may not be generalizable because persons were from an HIV centre of excellence, which may not be representative for smaller centres or primary care settings, social desirability bias, immortal time bias could have affected the study findings. Also, this is a non-randomized comparison and is subject to unmeasured confounding. Twenty percent of viral load data were missing but we used multiple imputation in the sensitivity analysis.

## Conclusion

The incidence of virological failure among PWA on long-term ART in this cohort study was low. Unlike the 30-day self-report, appointment keeping and 7-day self-reported ART adherence measures had low predictive performance in identifying virological failure. Routine plasma viral load monitoring remains the gold standard for adherence monitoring and confirming HIV treatment response.

## Data Availability

Data will be uploaded as a supplementary file if the manuscript is accepted for publication

## Acknowledgements

This project was supported by the Fogarty International Center of the National Institutes of Health (NIH) under Award Number D43TW009343 and the University of California Global Health Institute (UCGHI). The content is solely the responsibility of the authors and does not necessarily represent the official views of the NIH or UCGHI. BC was partly supported by the Fogarty International Centre, National Institute of Health (grant# 2D43TW009771-06 “HIV and co-infections in Uganda”).

## Author Contributions

**Conceptualization:** Stephen Okoboi, Barbara Castelnuovo, Rachel King

**Data curation:** Stephen Okoboi and Joseph Musaazi

**Formal analysis:** Stephen Okoboi, Joseph Musaazi, Izudi Jonathan, Sheri A. Lippman

**Methodology:** Stephen Okoboi, Joseph Musaazi, Rachel King, Sheri A. Lippman, Andrew Kambugu, Andrew Mujugira, Jonathan Izudi, Rosalind Parkes-Ratanshi, Agnes N. Kiragga, and Barbara Castelnuovo

**Project administration:** Stephen Okoboi

**Supervision:** Rachel King, Sheri A. Lippman and Barbara Castelnuovo

**Review and approval:** Stephen Okoboi, Joseph Musaazi, Rachel King, Sheri A. Lippman, Andrew Kambugu, Andrew Mujugira, Jonathan Izudi, Rosalind Parkes-Ratanshi, Agnes N. Kiragga, and Barbara Castelnuovo

